# Paired plasma and EV-enriched plasma proteomics reveal nonredundant sepsis-associated host-response signatures in critical illness

**DOI:** 10.64898/2026.06.11.26355454

**Authors:** SJ Rice, A Khaleghi Ardabili, V Ruiz-Velasco, AS Bonavia

## Abstract

**Background:** Plasma proteomics may identify host-response signatures in sepsis, but it is unclear whether extracellular vesicle (EV)-enriched plasma provides distinct or redundant information compared with plasma. We compared paired plasma and EV-enriched plasma proteomes in critically ill patients with sepsis and critically ill non-sepsis controls (CINS).

**Methods:** In this prospective observational study, paired plasma and EV-enriched plasma samples were analyzed from 56 critically ill adults, including 40 patients with sepsis and 16 CINS patients. Protein abundance was quantified using liquid chromatography-tandem mass spectrometry. Analyses compared proteomic depth, protein overlap, global concordance between compartments, and differential protein abundance between CINS and sepsis. Exploratory Gene Ontology enrichment was performed as a supplementary analysis.

**Results:** EV-enriched plasma expanded proteomic detection, identifying 2,476 filtered proteins compared with 506 in plasma. Only 386 proteins were detected in both compartments, while 2,090 were unique to EV-enriched plasma and 120 were unique to plasma. Among shared proteins, plasma and EV-enriched plasma showed modest global concordance across critically ill patients (Spearman ρ = 0.322, p = 9.19 x 10^-11^), with similar findings in sepsis alone. Differential abundance analysis identified 11 sepsis-associated proteins in plasma and 22 in EV-enriched plasma. Only SAA1, SAA2, and IGFBP6 were significant in both compartments. Exploratory pathway analysis supported acute-phase and inflammatory enrichment in plasma sepsis-associated proteins, while EV-enriched signals were directionally plausible but did not meet prespecified FDR thresholds.

**Conclusion:** Plasma and EV-enriched plasma proteomics capture related but nonredundant sepsis-associated host-response information in critically ill patients.

## Introduction

Extracellular vesicles (EVs) are increasingly recognized as biologically active participants in sepsis rather than passive circulating debris. Released by activated, injured, or apoptotic cells, EVs carry membrane and cargo proteins, lipids, and nucleic acids that can reflect the state of their parent cells and influence recipient-cell behavior (1). This biology is particularly relevant to sepsis, where leukocyte activation, endothelial injury, platelet activation, coagulation, and innate immune signaling converge. Consistent with this, circulating EVs have been implicated as both markers and mediators of immune dysregulation, endothelial injury, platelet activation, and immunothrombosis in sepsis (2, 3).

Clinical studies support the relevance of EVs in sepsis. Circulating EV features have been associated with sepsis outcomes (4), and EV-focused proteomic studies have identified sepsis-associated signatures involving innate immunity, complement and coagulation, platelet activation, endothelial activation, neutrophil pathways, and acute-phase biology (5, 6). These findings suggest that EV-enriched plasma may expose biologically informative protein signals that are less apparent in unfractionated plasma. At the same time, EV studies are technically heterogeneous, and current guidance emphasizes careful terminology and transparent reporting because EV-enriched preparations may contain overlapping vesicular and non-vesicular material (1). Thus, the clinical value of EV-enriched proteomics depends not only on the presence of EV-associated signals, but on whether they add information beyond standard plasma proteomics.

This distinction matters because plasma proteomics already captures clinically meaningful sepsis biology. High-throughput mass spectrometry has mapped host-response pathways, disease severity, organ dysfunction, and outcome-related proteomic features in sepsis (7). In our prior ICU cohort, plasma proteomics supported a parsimonious clinic-first model for sepsis recognition, reinforcing the value of circulating protein signatures in distinguishing sepsis from other critical illness (8). However, unfractionated plasma is dominated by abundant soluble proteins and may underrepresent cell-associated, membrane-associated, or vesicle-enriched signals. Whether EV-enriched plasma provides a distinct proteomic view of sepsis biology in the same patients remains uncertain.

Prior human sepsis EV proteomic studies have largely analyzed EVs or exosomes as standalone compartments, often using healthy comparator groups, rather than directly benchmarking EV-enriched plasma against paired unfractionated plasma (5, 6). To our knowledge, no prior study has specifically evaluated whether paired plasma and EV-enriched plasma provide redundant or complementary proteomic information in the clinically relevant comparison of sepsis versus noninfectious critical illness. This gap is important because EV enrichment may expand detectable proteomic depth while also introducing compartment-specific biology and pre-analytical complexity.

In the present study, we compared paired plasma and EV-enriched plasma proteomes from critically ill patients with sepsis and critically ill non-sepsis controls. We hypothesized that EV-enriched plasma would provide related but nonredundant proteomic information compared with plasma. The primary outcome was the degree of plasma-versus-EV-enriched plasma proteomic overlap and concordance, assessed by protein detection depth, shared protein abundance relationships, and compartment-specific sepsis-associated proteins.

## Materials and Methods

### Study Design and Participants

This prospective observational study was conducted at a quaternary academic medical center between November 2021 and August 2023 and is reported in accordance with STROBE guidelines. The study was approved by the institutional Human Subjects Protection Office, and informed consent was obtained from adult participants or their legally authorized representatives. Critically ill patients were identified through automated electronic health record screening using a Modified Early Warning Score algorithm, followed by independent manual review by two investigators to confirm eligibility.

Adults aged 18 years or older were eligible if enrolled within 48 h of critical illness onset. Sepsis was defined according to Sepsis-3 criteria as suspected or confirmed infection with an acute increase in SOFA score of at least 2 points (9). Critically ill non-sepsis (CINS) patients were enrolled using the same screening framework but did not meet Sepsis-3 criteria. Critical illness was defined by the requirement for continuous intravenous vasopressor support or noninvasive or invasive respiratory support. Patients were excluded for active hematologic malignancy, chronic immune-modifying therapy, chronic corticosteroid use unrelated to acute sepsis treatment, HIV infection with CD4 count <200 cells/mm³, chemotherapy or radiotherapy within 30 days, solid organ transplantation with ongoing immunosuppression, or autoimmune disease requiring immune-modifying treatment.

Clinical variables were abstracted from the electronic health record during hospitalization and supplemented by 30-day follow-up. Comorbidity burden was summarized using the Charlson Comorbidity Index, and acute illness severity was quantified using APACHE II. Shock was defined as serum lactate >2 mmol/L despite fluid resuscitation with >30 mL/kg intravenous crystalloid and ongoing vasopressor requirement. Stress-dose corticosteroid exposure was defined as receipt of at least 50 mg intravenous hydrocortisone for at least 8 h before blood sampling. Non-survivors were defined as participants who died within 30 days of enrollment.

### Blood Collection and Proteomic Sample Preparation

Blood for proteomic analysis was collected in K2-EDTA tubes and stored at 4°C immediately after collection. Plasma was isolated by centrifugation and stored at -80°C until\ batch processing. At the time of processing, plasma samples were thawed at 4°C and supplemented with protease inhibitor to limit protein degradation.

Proteomic profiling was performed on paired plasma and EV-enriched plasma samples from the same individuals. EV-enriched fractions were generated from plasma using a magnetic-particle-based positive selection workflow with a commercially available human pan-extracellular vesicle isolation kit (EasySep Cat# 17891, STEMCELL Technologies). This approach uses antibody-based capture of EVs expressing canonical tetraspanin surface markers, including CD9, CD63, and CD81. Enrichment was performed according to the manufacturer’s protocol, with parallel sample handling across batches to reduce technical variability. Because this approach preferentially enriches EV-associated material but does not produce purified vesicles, the resulting fraction is referred to throughout as EV-enriched plasma. Proteins from plasma and EV-enriched plasma fractions were extracted and digested using Trypsin/Lys-C under controlled digestion conditions before LC-MS/MS analysis.

### Liquid Chromatography-tandem mass spectrometry (LC-MS/MS) Proteomics

Proteomic analysis was performed using LC-MS on a Bruker timsTOF Flex instrument operated in diaPASEF mode. Peptides were separated by nanoflow liquid chromatography using reversed-phase C18 chromatography before data-independent acquisition. Instrument performance was monitored using regular calibration and quality-control digest runs. Protein identification and quantification were performed using a predicted spectral-library-based for the entire human proteome with DIANN software (10), generating protein-level abundance matrices for plasma and EV-enriched plasma samples.

Proteomic quality control included assessment of protein detection depth, missingness-based filtering, technical reproducibility, and detection of canonical EV-associated proteins. . In addition, yeast enolase protein was spiked into each sample following EV capture and trypsin-digested equine myoglobin was spiked into the samples following digestion cleanup to monitor proteomic processing and instrument performance throughout the experiment, respectively. For plasma proteomics, reproducibility was assessed by triplicate analysis of three samples from each clinical group and one control plasma sample; 90% of samples had a median coefficient of variation below 20%. Protein groups were retained after missingness filtering, followed by imputation.

### Proteomic Data Processing and Statistical Analysis

Plasma and EV-enriched plasma datasets were analyzed as paired but distinct proteomic compartments. Protein-level abundance matrices were filtered using the final post-processing outputs from the mass spectrometry workflow. Analyses were performed first to describe protein detection depth and overlap between compartments, then to evaluate concordance between paired plasma and EV-enriched plasma measurements among shared proteins.

The primary clinical contrast was sepsis versus critically ill non-sepsis. Protein groups were retained for analysis if present in at least 75% of samples within the corresponding dataset, followed by KNN imputation of missing values after filtering. Differential abundance was analyzed separately for plasma and EV-enriched plasma using the final post-processing outputs from the mass spectrometry workflow. Differentially abundant protein groups were defined by an absolute CINS-versus-sepsis log2 fold change ≥1 and Benjamini-Hochberg false-discovery rate (FDR) <0.05. Negative CINS-versus-sepsis log2 fold-change values were interpreted as higher abundance in sepsis, whereas positive values were interpreted as higher abundance in CINS.

Plasma SAA1 was measured by ELISA (Cat #KHA0011, Invitrogen) and evaluated as an orthogonal corroborative assay for the plasma proteomic SAA1 signal in available matched samples. Because ELISA was performed on plasma only, these data were not used to validate EV-enriched plasma proteomic measurements.

All analyses were designed to test whether plasma and EV-enriched plasma provide related but nonredundant proteomic information in critical illness and sepsis. Accordingly, pathway-level analyses were treated as exploratory and were not used as primary evidence for biological mechanism.

## Results

### Sepsis and CINS Patients Represented Clinically Comparable Critically Ill Cohorts

The primary analysis compared critically ill patients with sepsis to CINS controls. Clinical characteristics, illness severity, organ support, shock status, corticosteroid exposure, and 30-day mortality are summarized in **Table 1**. This comparison was selected to evaluate whether plasma and EV-enriched plasma proteomics provide complementary information within the clinically relevant ICU setting, where both groups have substantial acute illness burden but differ by the presence or absence of sepsis.

**Table 1.** Clinical characteristics of critically ill patients included in the paired proteomics analysis. Values are summarized by clinical group among the 56 critically ill patients with matched plasma and EV-enriched plasma proteomics. Continuous variables are reported as median [IQR], and binary variables as n/N (%). *P-*values are exploratory and were calculated using Mann-Whitney U tests for continuous variables and Fisher exact or chi-square tests for binary variables.

| Characteristic | CINS (n=16) | Sepsis (n=40) | *P*-value |
| --- | --- | --- | --- |
| Age, years | 68.5 [60.8, 75.2] | 71.0 [59.8, 77.5] | 0.586 |
| BMI, kg/m<sup>2</sup> | 26.0 [22.1, 31.1] | 31.2 [26.0, 44.8] | 0.015 |
| APACHE II | 22.5 [15.8, 26.2] | 22 [16, 31] | 0.388 |
| SOFA day 1 | 7.5 [5.8, 10.2] | 8.0 [7.0, 11.2] | 0.471 |
| Maximum SOFA | 9.5 [5.8, 11.0] | 8.0 [7.0, 11.2] | 0.935 |
| Lactate, mmol/L | 2.8 [1.7, 4.9] | 3.1 [2.1, 4.8] | 0.797 |
| Creatinine, mg/dL | 1.0 [0.8, 1.3] | 1.8 [1.3, 2.6] | 0.001 |
| BUN, mg/dL | 17.0 [14.8, 21.2] | 37.0 [24.8, 51.0] | <0.001 |
| Platelets, x10<sup>3</sup>/uL | 175.0 [135.5, 206.8] | 157.0 [100.2, 199.0] | 0.318 |
| Length of stay, days | 11.0 [3.8, 20.5] | 8.0 [5.8, 12.2] | 0.716 |
| Male | 9/16 (56%) | 23/40 (58%) | 0.932 |
| Shock | 6/16 (38%) | 23/40 (58%) | 0.176 |
| Mechanical ventilation | 10/15 (67%) | 13/40 (32%) | 0.022 |
| Bacteremia | 0/16 (0%) | 24/40 (60%) | <0.001 |
| In-hospital mortality | 3/16 (19%) | 2/40 (5%) | 0.135 |
| 30-day mortality | 3/16 (19%) | 3/40 (8%) | 0.338 |
| 90-day mortality | 5/16 (31%) | 7/40 (18%) | 0.257 |
| Corticosteroid use | 4/16 (25%) | 8/40 (20%) | 0.726 |

Primary hospital diagnoses for CINS patients are shown in Supplementary Table 1. Among patients with sepsis, most had culture-positive infection (93%). Gram-negative infection was identified in 21 patients (53%), while 13 patients (33%) had gram-positive or mixed gram-positive/gram-negative infection. The remaining patients had fungal or viral infection.

### EV-Enriched Plasma Provided Greater Proteomic Depth and Partial Overlap with Plasma

In the EV-enriched plasma dataset, canonical EV-associated proteins, including CD9, CD63, and CD81, were detected and did not differ significantly across clinical groups. EV-enriched plasma yielded substantially greater proteomic depth than plasma. After filtering, 2,476 proteins were identified in EV-enriched plasma compared with 506 proteins in plasma (**Fig 1**; **Table 2**). Among these, 386 proteins were detected in both compartments, while 2,090 proteins were detected only in EV-enriched plasma and 120 only in plasma. Thus, EV-enriched plasma expanded detectable protein coverage but did not simply reproduce the plasma proteome.

**Figure 1.**
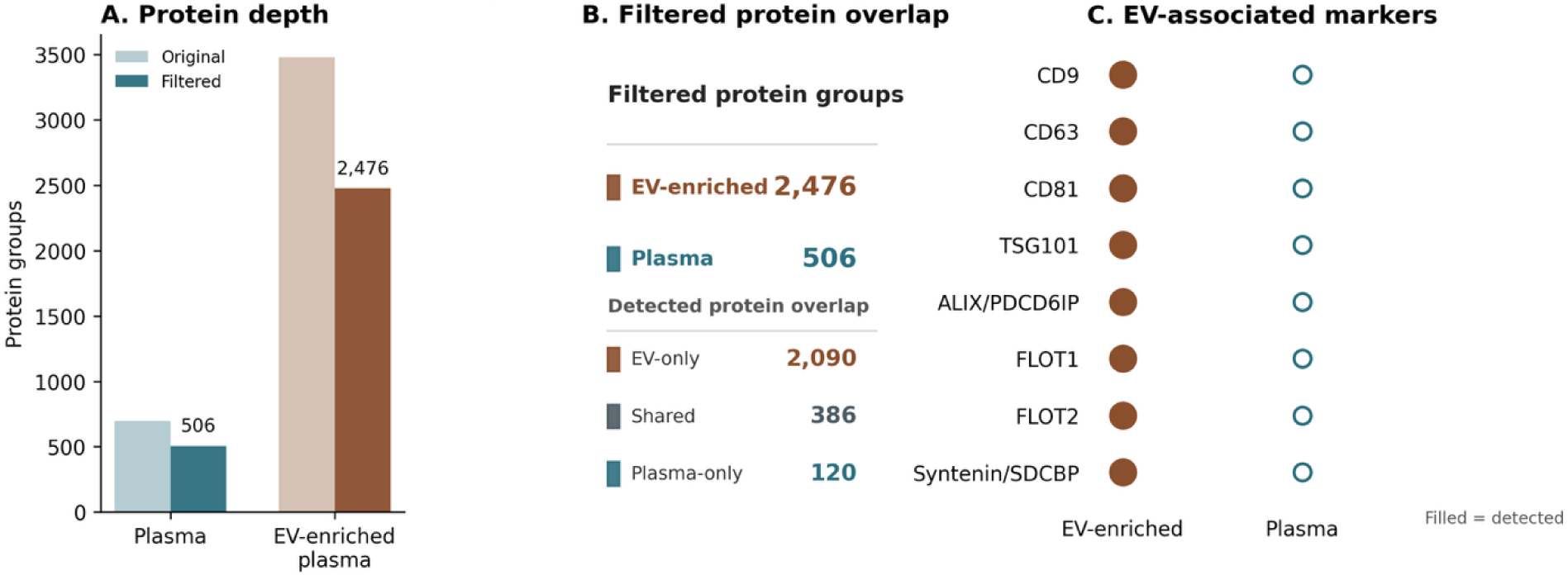
Paired plasma and EV-enriched plasma proteomic datasets compare distinct circulating fractions. Matched plasma and EV-enriched plasma samples from the same cohort were analyzed by LC-MS-based proteomics. In the EV-enriched plasma dataset, canonical EV-associated proteins, including CD9, CD63, and CD81 were detected and did not differ significantly across clinical groups. **A**, Protein group depth before and after filtering was greater in EV-enriched plasma than in plasma. **B**, Among filtered protein groups, only a subset was detected in both compartments, with most detected proteins unique to the EV-enriched plasma dataset. **C**, Canonical EV-associated proteins were detected in the EV-enriched plasma dataset, supporting enrichment of vesicle-associated material.

**Table 2.**
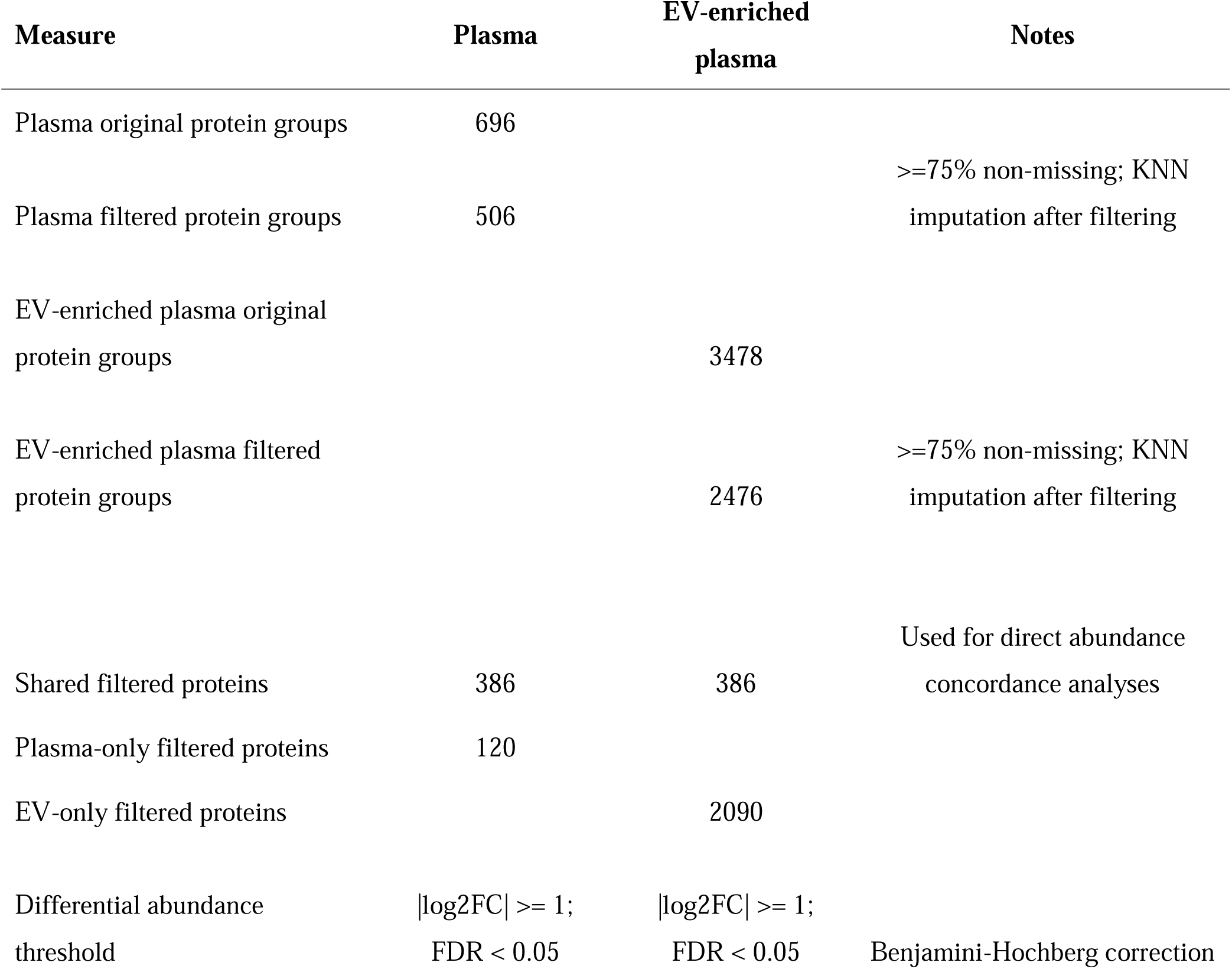
Summary of paired plasma and EV-enriched plasma proteomics datasets. This table summarizes sample counts, protein group depth, filtering, overlap, and differential abundance thresholds for the paired proteomic datasets.

### Plasma and EV-Enriched Plasma were Related but not Strongly Concordant

Across critically ill patients, the global plasma-versus-EV-enriched plasma protein-level correlation was modest (Spearman ρ = 0.322, p = 9.19 x 10^-11^; Pearson r = 0.372; **Fig 2**). Similar results were observed when restricted to patients with sepsis (Spearman ρ = 0.294, p = 3.76 x 10^-9^; Pearson r = 0.350). These findings indicate that paired plasma and EV-enriched plasma proteomics capture related but nonredundant protein abundance patterns.

**Figure 2.**
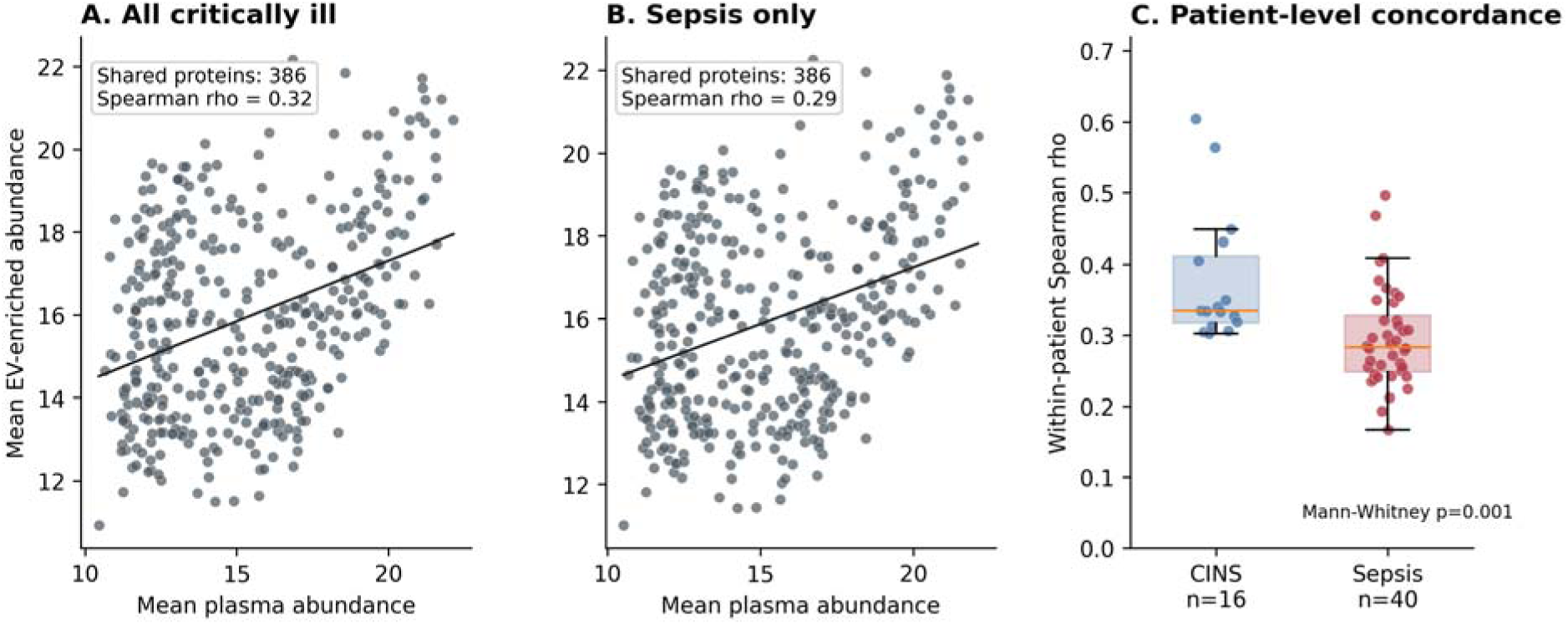
Global plasma and EV-enriched plasma protein abundance patterns are modestly concordant. Among the 386 proteins detected in both compartments, mean protein abundance patterns were compared between plasma and EV-enriched plasma. **A**, In all critically ill patients, global abundance concordance between compartments was modest. **B**, Similar concordance was observed when the analysis was restricted to patients with sepsis. **C**, Patient-level plasma/EV-enriched plasma concordance varied across individuals and was lower in sepsis than in CINS in this exploratory comparison. These findings support the interpretation that EV-enriched plasma is not simply a proportional representation of the plasma proteome.

### Sepsis-Associated Protein Differences were mostly Compartment-Specific

Differential abundance analysis comparing CINS versus sepsis identified 11 significant proteins in plasma and 22 significant proteins in EV-enriched plasma (**Fig 3**; **Supplementary Tables 2 - 4**). Most significant proteins were unique to one compartment. Plasma significant proteins included CRP, LBP, LCN2, SAA1, SAA2, VWF, B2M, THBS4, IGFBP6, LILRA3, and B4GALT5. EV-enriched plasma significant proteins included EBI3, PLA2G2A, IGF1, MGP, MMP14, ELN, SERPINC1, LTBP2, ADAMTS5, SPON2, ALB, TF, B4GALT1, IGFBP6, SAA2, MFGE8, SAA1, RBP4, TRAP1, PLSCR1, CHRD, and PCSK9.

**Figure 3.**
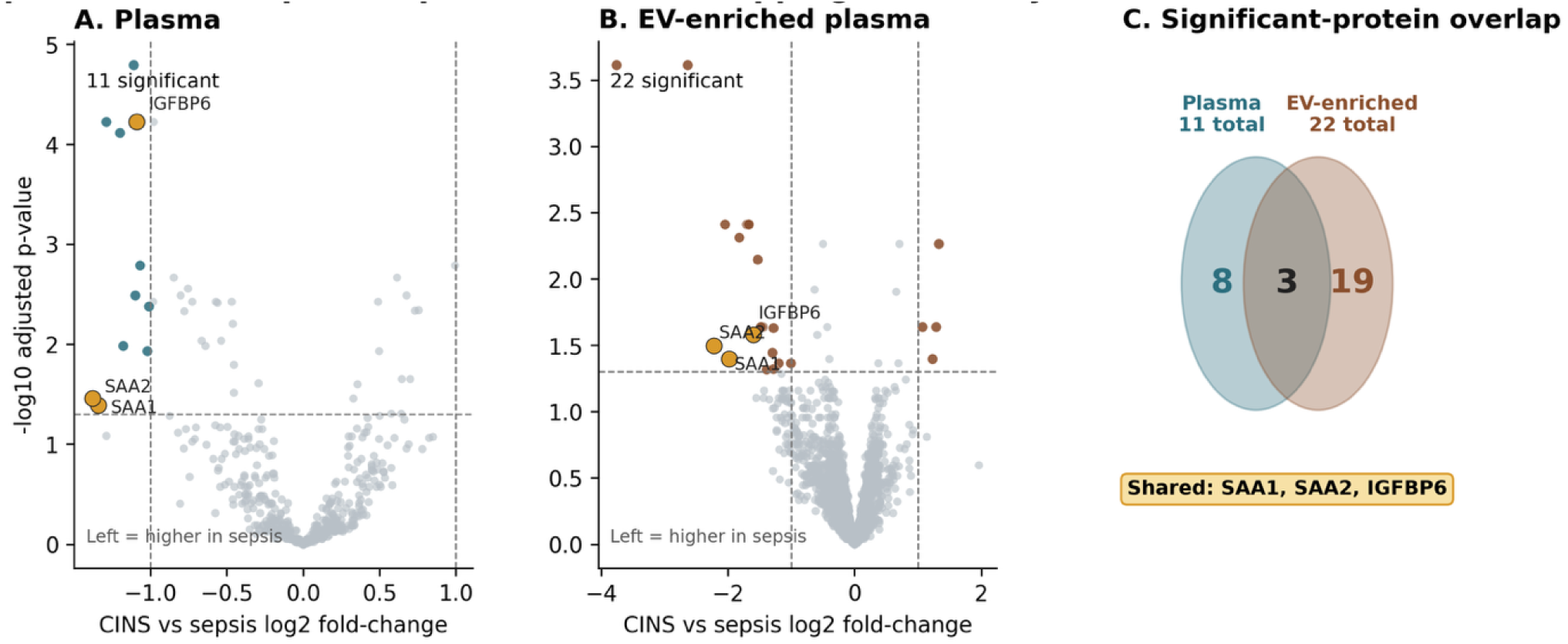
Sepsis-associated protein patterns are overlapping but mostly nonredundant between plasma and EV-enriched plasma. Differential abundance analysis comparing critically ill non-sepsis patients with sepsis patients was performed separately in plasma and EV-enriched plasma. **A**, Volcano plot of sepsis-associated proteins in plasma. **B**, Volcano plot of sepsis-associated proteins in EV-enriched plasma. Negative CINS-vs-sepsis log2 fold-change indicates higher abundance in sepsis. **C**, Overlap of significant proteins between compartments. Most significant proteins were compartment-specific, while SAA1, SAA2, and IGFBP6 were significant in both plasma and EV-enriched plasma.

Only three proteins were significant in both compartments: serum amyloid A1 (SAA1), SAA2, and insulin-like growth factor-binding protein 6 (IGFBP6) (**Table 3**). Therefore, while both plasma and EV-enriched plasma captured sepsis-associated biology, most differentially abundant proteins were compartment-specific, supporting the premise that EV-enriched plasma and plasma provide complementary rather than interchangeable proteomic information.

**Table 3.**
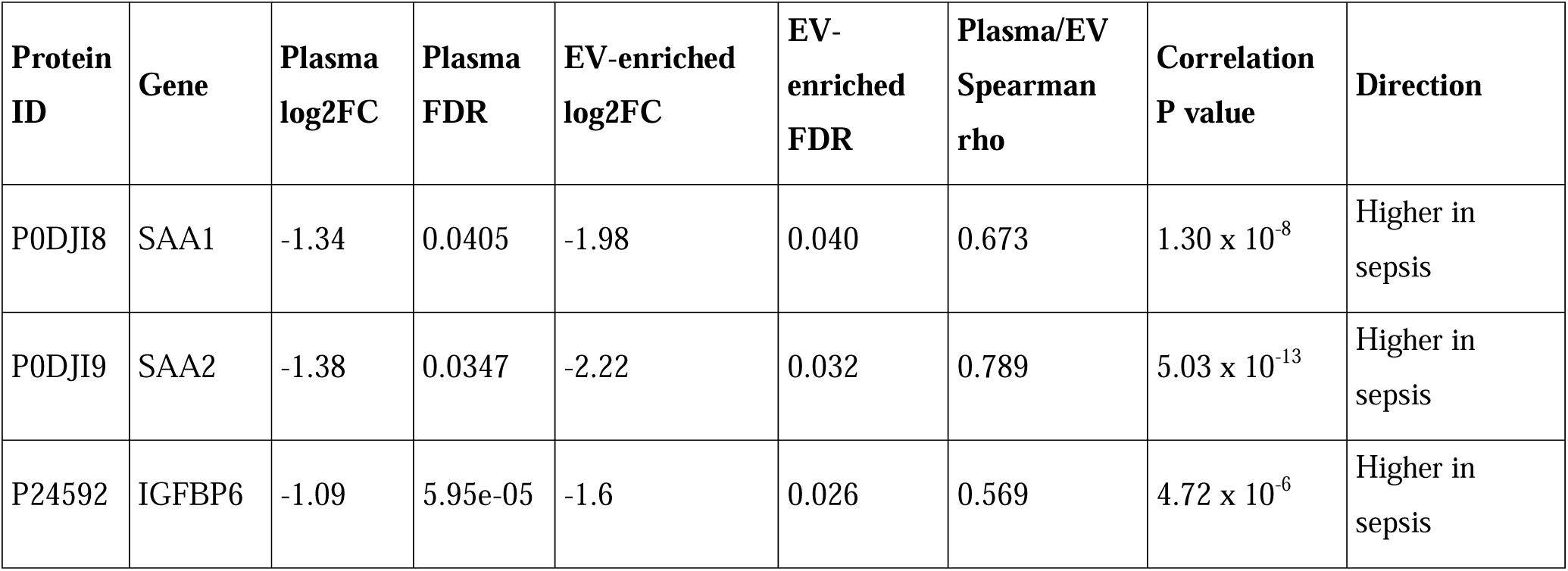
Shared sepsis-associated proteins detected in both plasma and EV-enriched plasma. This table lists proteins significantly different between CINS and sepsis in both compartments, including compartment-specific log2 fold-changes, FDR values, and matched-sample plasma/EV-enriched plasma Spearman correlations.

### Shared Sepsis-Associated Proteins Retained Positive Cross-Compartment Relationships

For the three proteins significant in both compartments, paired plasma and EV-enriched plasma abundance values showed positive correlations, although the strength of association varied by protein (**Fig 4**; **Table 3**). In critically ill patients, Spearman correlations were strongest for SAA2 (ρ = 0.789), followed by SAA1 (ρ = 0.673) and IGFBP6 (ρ = 0.569). In the sepsis subgroup, correlations remained positive but were lower for SAA2 and SAA1. These results suggest that shared sepsis-associated proteins retain related signal across compartments, while still leaving substantial room for compartment-specific information.

**Figure 4.**
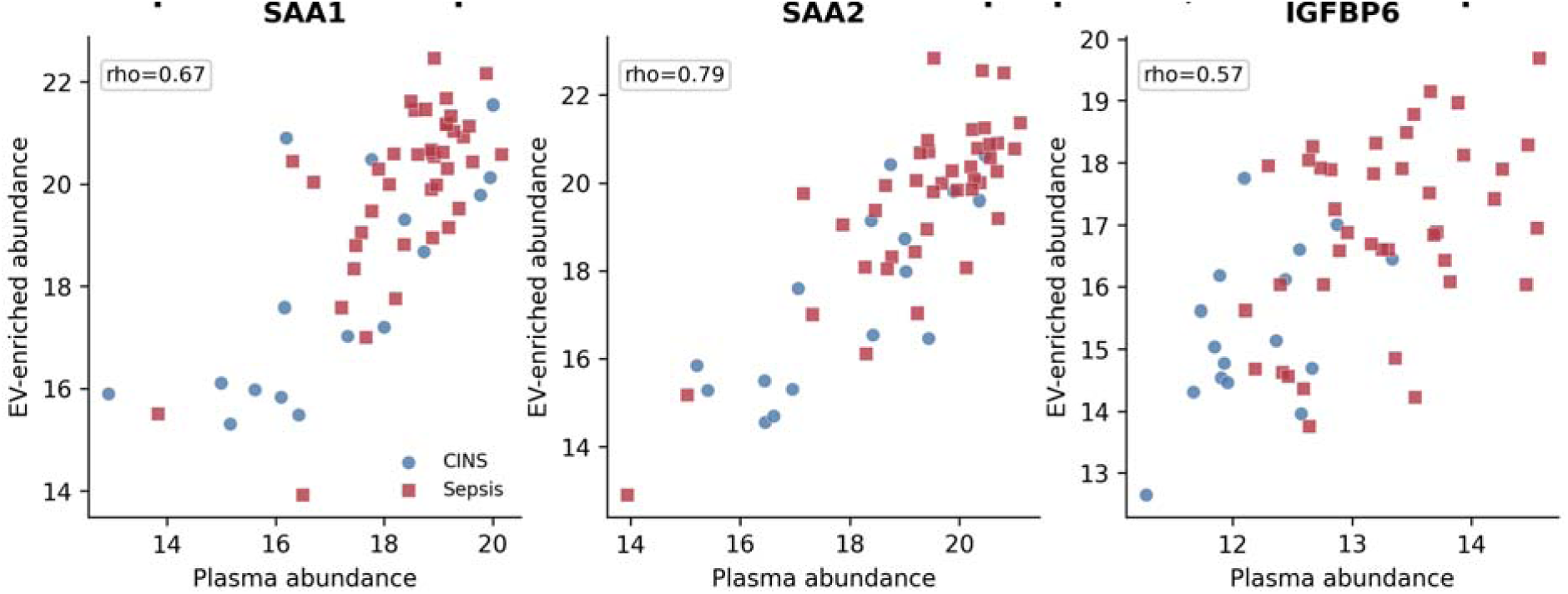
Shared sepsis-associated proteins show matched-sample plasma/EV-enriched plasma relationships. SAA1, SAA2, and IGFBP6 were the only proteins significantly associated with sepsis in both plasma and EV-enriched plasma. Matched-sample abundance plots demonstrate moderate to strong cross-compartment relationships, particularly for SAA1 and SAA2. These shared proteins may represent robust systemic sepsis-associated signals detectable across circulating protein compartments.

Plasma SAA1 measured by ELISA was available for orthogonal comparison with plasma proteomic SAA1. After excluding non-computable (out of reliable measurable range) values, 47 matched samples were analyzed. Plasma ELISA SAA1 correlated with plasma proteomic SAA1 abundance (Spearman ρ = 0.56, p = 3.78 x 10^-5^; **Supplementary Fig. 1**). Because ELISA was performed on plasma only, this analysis was interpreted as corroboration of the plasma SAA1 proteomic signal and not as validation of EV-enriched plasma measurements.

### Exploratory Pathway Enrichment Supported Acute-Phase and Inflammatory Biology

Exploratory GO Biological Process enrichment was performed using the final CINS-versus-sepsis protein lists, a common tested-protein background, and Benjamini-Hochberg correction (**Supplementary Fig 2**). Plasma proteins higher in sepsis showed enrichment for acute-phase response, acute inflammatory response, bacterial defense-response terms, and neutrophil or granulocyte chemotaxis/migration. EV-enriched plasma proteins higher in sepsis showed directionally plausible terms, including acute inflammatory response, acute-phase response, macrophage migration, cell adhesion, and leukocyte activation, but these did not meet FDR < 0.10.

## Discussion

In this paired proteomic analysis of critically ill patients with sepsis and critically ill non-sepsis controls, EV-enriched plasma substantially expanded detectable protein depth compared with plasma, but the two compartments overlapped only partially and showed modest global concordance. Differential abundance analysis identified sepsis-associated proteins in both compartments, yet most significant proteins were compartment-specific. Only SAA1, SAA2, and IGFBP6 were significant in both plasma and EV-enriched plasma. Together, these findings support the central premise that EV-enriched plasma proteomics provides a circulating host-response readout that is related to, but not interchangeable with, standard plasma proteomics.

These results extend prior plasma proteomic studies in sepsis. Large-scale mass-spectrometry studies have shown that the plasma proteome captures biologically coherent sepsis pathways, including inflammation, complement activation, coagulation, organ dysfunction, disease severity, clinical subphenotypes, and mortality-associated trajectories (7, 11). Our prior work similarly demonstrated that plasma proteomics can support a parsimonious, clinic-first model for sepsis recognition in the ICU (8). The present study builds on that foundation by asking a narrower question: whether EV-enriched plasma provides additional or redundant information when compared directly with paired plasma from the same critically ill patients. The modest global concordance observed here suggests that EV enrichment does not simply rescale or concentrate the plasma proteome but instead changes the observable protein space.

Our findings are also consistent with emerging human EV studies in sepsis, while addressing a different clinical comparison. Morris et al. reported that plasma-derived exosome proteomic profiles distinguished septic emergency department patients from healthy controls, with acute-phase proteins including SAA1, SAA2, and C-reactive protein (CRP) among the most prominent differences (5). More recently, Park et al. performed plasma EV proteomics in septic patients and healthy controls and identified signatures involving innate immunity, complement and coagulation, platelet activation, endothelial activation, neutrophil extracellular trap formation, and inflammatory pathways (6). In our study, SAA1 and SAA2 again emerged as robust sepsis-associated proteins, supporting the reproducibility of acute-phase biology across proteomic platforms and EV-enrichment strategies. However, unlike prior healthy-control comparisons, our primary contrast was sepsis versus noninfectious critical illness. This comparison is clinically more stringent because both groups share organ dysfunction, ICU-level physiology, and systemic inflammation. The smaller number of significant proteins and the limited pathway enrichment in EV-enriched plasma likely reflect this more difficult comparator rather than absence of biological signal.

The paired design is an important distinction from much of the existing EV literature. Many EV studies analyze EVs or exosomes as standalone biomarker compartments, but do not determine whether those measurements add information beyond paired unfractionated plasma. Work outside human sepsis has shown that soluble plasma and plasma EV protein compartments are overlapping but distinct, supporting the concept that EV-enriched fractions can expand the observable proteome (12). A recent murine bacterial sepsis study similarly found that EV and plasma proteomes were altered differently during systemic infection, with EVs emphasizing cellular processes and signaling while plasma reflected other systemic protein changes (13). Our human ICU data are concordant with this concept: EV-enriched plasma identified many more proteins than plasma and included compartment-specific sepsis-associated proteins but retained only modest cross-compartment concordance.

The shared proteins are also informative. SAA1 and SAA2 are canonical acute-phase proteins (14, 15) and were significant in both compartments, with positive plasma/EV-enriched plasma relationships. Plasma SAA1 measured by ELISA further corroborated the plasma proteomic SAA1 signal, although the ELISA data were semi-quantitative because many samples remained outside the preferred optical-density range despite 9000-fold dilution. These findings support SAA1 as a robust systemic sepsis-associated signal (16). This interpretation is consistent with prior sepsis EV proteomic studies in which acute-phase proteins, including SAA1 and SAA2, were among the most prominent inflammatory signals (5, 6). At the same time, the fact that SAA1 and SAA2 were among the few shared proteins highlights an important point: strong systemic acute-phase signals may be detectable across circulating compartments, whereas many other proteins appear more compartment-restricted. EV-associated acute-phase biology has also been described for CRP-positive EVs in sepsis, reinforcing that classical soluble inflammatory proteins may interact with, bind to, or co-isolate with EV-associated material (17).

These observations have practical implications for biomarker discovery. EV-enriched plasma may be useful not because it replaces plasma, but because it broadens discovery space and may reveal cell-associated, membrane-associated, immune, endothelial, coagulation, or extracellular-matrix signals that are less apparent in unfractionated plasma. For clinical sepsis research, this distinction matters. A biomarker that distinguishes sepsis from health may have limited bedside relevance; a biomarker compartment that adds information within critically ill patients with overlapping clinical syndromes is potentially more useful. The present findings suggest that EV-enriched plasma could complement plasma proteomics in future sepsis phenotyping studies, especially if paired designs are used to distinguish robust systemic signals from compartment-specific biology.

Several limitations should temper interpretation. First, this was a modest-sized, single-center cohort, and findings require validation in independent critically ill populations. Second, critically ill non-sepsis controls are clinically heterogeneous, and residual diagnostic misclassification between infectious and noninfectious critical illness is possible despite adjudication. Third, EV-enriched plasma should not be interpreted as purified EV cargo. Current MISEV guidance emphasizes transparent terminology because EV preparations may include vesicular and non-vesicular extracellular particles, lipoproteins, soluble proteins, protein aggregates, and proteins bound to the EV surface or corona (1). Plasma is particularly challenging for EV proteomics because abundant plasma proteins and lipoproteins can co-isolate with EV fractions (18). This concern is especially relevant to immunocapture-based workflows, which enrich EV subpopulations based on surface markers but may not capture all EVs and may still recover associated non-vesicular material (19). Fourth, this analysis was cross-sectional and does not address temporal changes in plasma or EV-enriched plasma proteomes during sepsis evolution or recovery. Finally, pathway enrichment and ELISA analyses were exploratory; they support biological plausibility but should not be treated as mechanistic proof.

In summary, paired plasma and EV-enriched plasma proteomics provide complementary views of the sepsis-associated host response in critical illness. EV-enriched plasma greatly expanded protein detection and revealed largely compartment-specific sepsis-associated proteins, while shared signals such as SAA1 and SAA2 reflected robust systemic acute-phase biology.

These findings support EV-enriched plasma as a complementary proteomic compartment for sepsis discovery studies, while underscoring the need for rigorous terminology, careful paired benchmarking against plasma, and independent validation before clinical application.

## Declarations

### Ethics approval and consent to participate

This study was conducted in accordance with the principles of the Declaration of Helsinki and was approved by the Penn State College of Medicine Institutional Review Board / Human Subjects Protection Office (IRB #15328). Written informed consent was obtained from adult participants or their legally authorized representatives, as appropriate, before enrollment. All data and biospecimens were de-identified before analysis to protect participant confidentiality.

### Consent for publication

Not applicable.

### Availability of data and materials

The data supporting the conclusions of this article are included within the article and its supplementary information files. Additional de-identified data generated or analyzed during the current study may be available from the corresponding author on reasonable request, subject to applicable institutional and ethical approvals.

### Authors’ contributions

ASB conceived and designed the study. SJR and VRV contributed to data acquisition. SJR, AKA, and ASB performed data analysis and interpretation. SJR and ASB drafted the manuscript. All authors critically revised the manuscript for important intellectual content. All authors read and approved the final manuscript and agree to be accountable for all aspects of the work.

## Data Availability

All data produced in the present study are available upon reasonable request to the authors

## Acknowledgements

The authors thank Ruth-Ann Brown and Kristen Brandt for their assistance with sample collection and processing. The authors also acknowledge the Mass Spectrometry and Proteomics Core (RRID:SCR_017831) for proteomics services and instrumentation support.

