## Supplementary Materials for "Paired plasma and EV-enriched plasma proteomics reveal nonredundant sepsis-associated host-response signatures in critical illness"

**SUPPLEMENTARY METHODS**

**LC-MS/MS Acquisition Details**

Tryptic digests were injected onto a PepMap Neo C18 trap column and separated on a 15 cm nanoflow C18 analytical column maintained at 50°C. The NanoElute2 system was operated at 500 bar with a flow rate of 1 µL/min. Mobile phases consisted of water with 0.1% formic acid and acetonitrile with 0.1% formic acid. The gradient began at 2% organic phase, increased to 30%, then to 90%, followed by wash and re-equilibration.

Data were acquired using a CaptiveSpray nano-electrospray source with capillary voltage of 1400 V, dry gas flow of 3.0 L/min, and dry temperature of 180°C. DIA acquisition used a diaPASEF method with MS1 scanning from 100–1700 m/z and ion mobility scanning from 0.60–1.60 1/K0. MS2 scans used consecutive non-overlapping isolation windows across the 400–1000 m/z range. Ion mobility calibration was performed using Agilent tune mix ions.

**Plasma SAA1 ELISA Corroboration**

Plasma SAA1 concentrations were measured by ELISA in available samples using plasma diluted 9000-fold. Mean optical density values were converted to estimated plasma SAA1 concentrations using the four-parameter logistic standard-curve parameters recorded with the assay and then multiplied by the dilution factor. One duplicated sample identifier with discordant ELISA optical density values was excluded from the primary analysis. Because many samples remained outside the preferred optical-density range despite 9000-fold dilution, reconstructed concentrations were treated as semi-quantitative. Associations between ELISA-derived plasma SAA1 and plasma proteomic SAA1 abundance were assessed using Spearman correlation, with a sensitivity analysis restricted to samples within the preferred optical-density range.

**Exploratory Pathway Enrichment**

As an exploratory supplementary analysis, Gene Ontology Biological Process over-representation analysis was performed using the final CINS-versus-sepsis significant protein lists from plasma and EV-enriched plasma. Protein lists were stratified by direction of effect. The same background universe was used for all enrichment tests, defined as the union of all gene symbols tested in either plasma or EV-enriched plasma and successfully mapped to Entrez identifiers. Enrichment was performed using *clusterProfiler* with org.Hs.eg.db, and p values were adjusted within each directional analysis using the Benjamini-Hochberg method. Because the significant protein lists were small, GO terms with a minimum size of two genes were permitted, and results were interpreted as hypothesis-generating only.

**SUPPLEMENTARY FIGURES**

**Supplementary Figure 1. Plasma SAA1 ELISA corroborates plasma proteomic SAA1 abundance.**
Plasma SAA1 measured by ELISA was compared with plasma proteomic SAA1 abundance in matched samples. After excluding one duplicated ambiguous sample and out-of-range values, 47 samples were included. Plasma ELISA SAA1 correlated with plasma proteomic SAA1 abundance (Spearman ρ = 0.56, p = 3.78 x 10^-5^). In the subset of samples within the preferred ELISA optical-density range, the correlation was stronger (Spearman ρ = 0.88, p = 0.0016; n = 9). Because many samples remained above the preferred optical-density range at the 9000-fold dilution, the ELISA results were interpreted as semi-quantitative corroboration rather than precise absolute concentration validation. ELISA was performed on plasma only and was not used to validate EV-enriched plasma proteomic measurements.

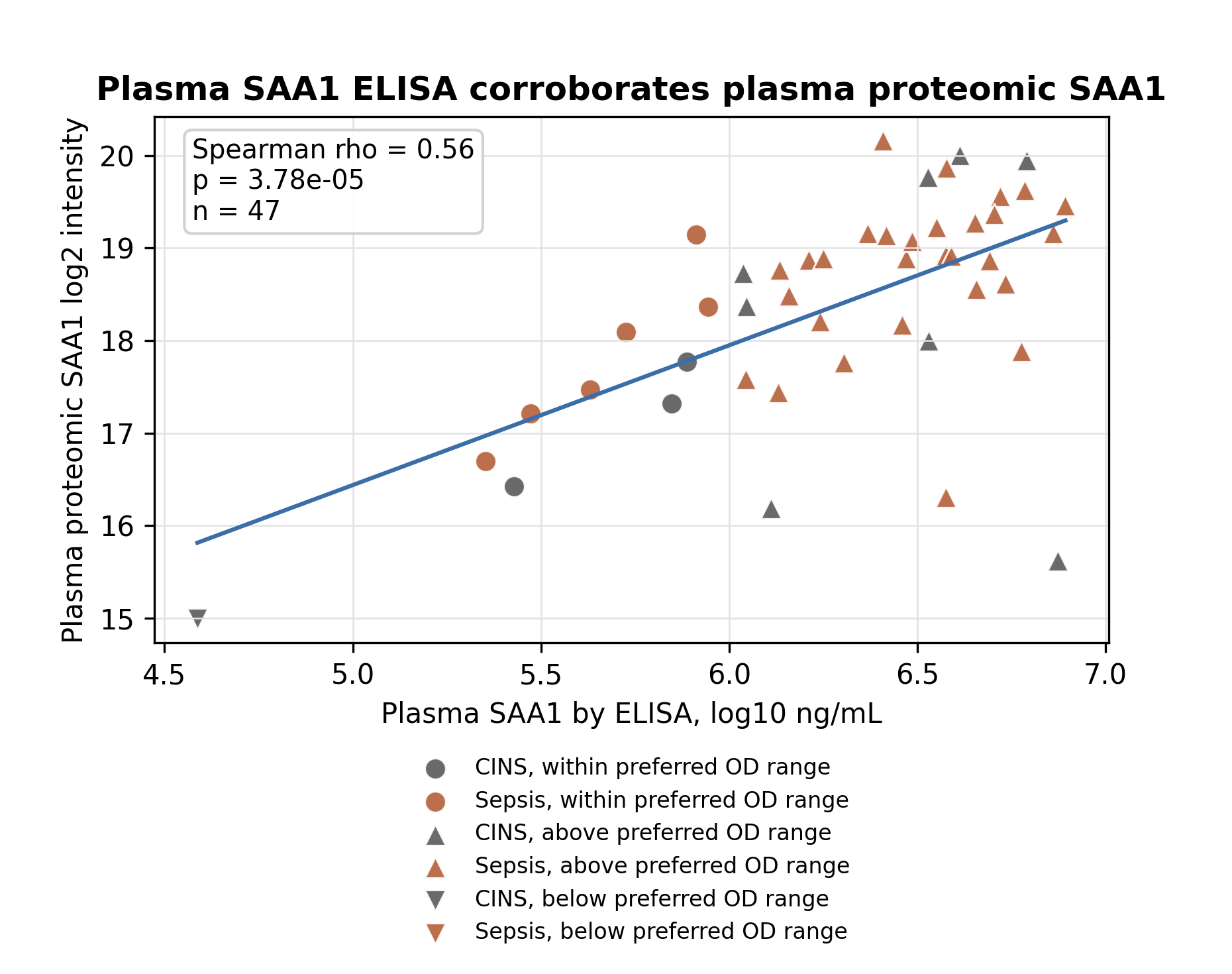

**Supplementary Figure 2. Exploratory GO Biological Process enrichment for CINS versus sepsis.**

Over-representation analysis was performed using final CINS-versus-sepsis significant proteins stratified by direction of effect in plasma and EV-enriched plasma. The same background universe, consisting of all genes tested in either compartment and mapped to Entrez identifiers, was used for all analyses. Points show top multi-gene GO Biological Process terms ranked by Benjamini-Hochberg-adjusted p value; the dashed line indicates FDR = 0.10. Plasma sepsis-high proteins showed enrichment for acute-phase, acute inflammatory, bacterial response, and neutrophil/granulocyte migration terms. EV-enriched plasma terms were directionally plausible but did not meet FDR < 0.10 in this exploratory analysis.

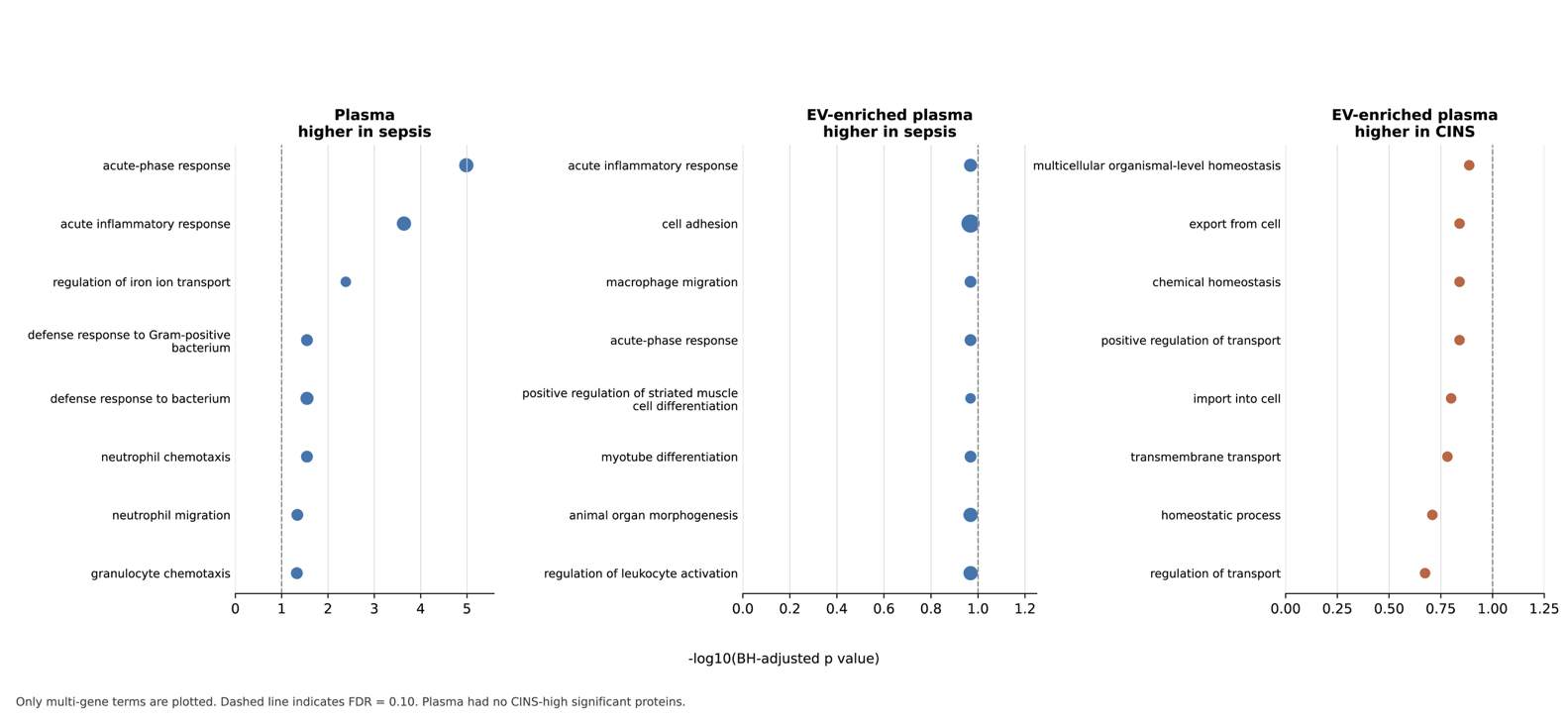

**SUPPLEMENTARY TABLES**

**Supplementary Table 1. Primary Hospital Diagnoses of Critically Ill Non-Sepsis Patients.**

| **CINS Patient Number** | **Primary Diagnosis** |
| --- | --- |
| 1 | Pulmonary edema and non-ST segment elevation myocardial infarction |
| 2 | Cardiogenic Shock |
| 3 | Coronary artery disease |
| 4 | Unilateral weakness, thrombocytopenia |
| 5 | Acute hypoxemic respiratory failure |
| 6 | Subarachnoid hemorrhage |
| 7 | Acute trauma |
| 8 | Acute pancreatitis |
| 9 | Traumatic Brain Injury |
| 10 | Post-operative stroke |
| 11 | Trauma, motor vehicle collision |
| 12 | Cirrhosis and shock |
| 13 | Cardiac arrest |
| 14 | Upper gastrointestinal bleeding |
| 15 | Trauma, motor vehicle collision |
| 16 | Seizure with Acute Respiratory Failure |

**Supplementary Table 2. Plasma CINS-vs-sepsis significant proteins.**

Full list of proteins significantly different between CINS and sepsis in plasma.

| **Protein ID** | **Gene Name** | **CINS vs sepsis log2FC** | **P-value** | **FDR** |
| --- | --- | --- | --- | --- |
| Q8N6C8 | LILRA3 | -1.11 | 3.21 x 10^-8^ | 1.62 x 10^-5^ |
| P24592 | IGFBP6 | -1.09 | 2.47 x 10^-7^ | 5.95 x 10^-5^ |
| P80188 | LCN2 | -1.29 | 4.3 x 10^-7^ | 5.95 x 10^-5^ |
| O43286 | B4GALT5 | -1.2 | 7.57 x 10^-7^ | 7.66 x 10^-5^ |
| P61769 | B2M | -1.07 | 2.24 x 10^-5^ | 0.00162 |
| P35443 | THBS4 | -1.1 | 7.68 x 10^-5^ | 0.00323 |
| P18428 | LBP | -1.01 | 0.000165 | 0.00419 |
| P02741 | CRP | -1.18 | 0.000578 | 0.0104 |
| P04275 | VWF | -1.02 | 0.000696 | 0.0117 |
| P0DJI9 | SAA2 | -1.38 | 0.00261 | 0.0347 |
| P0DJI8 | SAA1 | -1.34 | 0.00312 | 0.0405 |

**Supplementary Table 3. EV-enriched plasma CINS-vs-sepsis significant proteins.**

Full list of proteins significantly different between CINS and sepsis in EV-enriched plasma.

| **Protein ID** | **Gene Name** | **CINS vs sepsis log2FC** | **P-value** | **FDR** |
| --- | --- | --- | --- | --- |
| Q14213 | EBI3 | -2.64 | 1.6 x 10^-7^ | 0.000244 |
| P14555 | PLA2G2A | -3.76 | 1.97 x 10^-7^ | 0.000244 |
| P05019 | IGF1 | -2.05 | 6.88 x 10^-6^ | 0.00388 |
| P08493 | MGP | -1.7 | 6.01 x 10^-6^ | 0.00388 |
| P50281 | MMP14 | -1.67 | 7.84 x 10^-6^ | 0.00388 |
| P15502 | ELN | -1.82 | 1.17 x 10^-5^ | 0.00484 |
| P01008 | SERPINC1 | 1.33 | 1.88e x 10^-5^ | 0.00541 |
| Q14767 | LTBP2 | -1.53 | 2.88 x 10^-5^ | 0.00712 |
| Q9UNA0 | ADAMTS5 | -1.48 | 0.000149 | 0.0229 |
| Q9BUD6 | SPON2 | -1.46 | 0.000123 | 0.0229 |
| P02768 | ALB | 1.07 | 0.000152 | 0.0229 |
| P02787 | TF | 1.29 | 0.000157 | 0.0229 |
| P15291 | B4GALT1 | -1.28 | 0.00017 | 0.0234 |
| P24592 | IGFBP6 | -1.6 | 0.000212 | 0.0262 |
| P0DJI9 | SAA2 | -2.22 | 0.00027 | 0.0319 |
| Q08431 | MFGE8 | -1.3 | 0.000318 | 0.0358 |
| P0DJI8 | SAA1 | -1.98 | 0.000377 | 0.04 |
| P02753 | RBP4 | 1.23 | 0.000404 | 0.04 |
| Q12931 | TRAP1 | -1.2 | 0.000465 | 0.0429 |
| O15162 | PLSCR1 | -1.01 | 0.000491 | 0.0429 |
| Q9H2X0 | CHRD | -1.28 | 0.000576 | 0.0476 |
| Q8NBP7 | PCSK9 | -1.39 | 0.0006 | 0.0479 |

**Supplementary Table 4. Compartment-specific and shared sepsis-associated proteins.**

Combined table of all proteins significant in plasma and/or EV-enriched plasma, annotated as plasma-only, EV-only, or shared.

| **Protein ID** | **Gene** | **Sig. in plasma** | **Sig. in EV-enriched** | **Compartment category** | **Plasma log2FC** | **Plasma FDR** | **EV-enriched log2FC** | **EV-enriched FDR** |
| --- | --- | --- | --- | --- | --- | --- | --- | --- |
| O15162 | PLSCR1 | False | True | EV-only |  |  | -1.01 | 0.0429 |
| O43286 | B4GALT5 | True | False | Plasma-only | -1.2 | 7.66 x 10^-5^ |  |  |
| P01008 | SERPINC1 | False | True | EV-only | 0.0921 | 0.698 | 1.33 | 0.00541 |
| P02741 | CRP | True | False | Plasma-only | -1.18 | 0.0104 | -0.479 | 0.472 |
| P02753 | RBP4 | False | True | EV-only | 0.479 | 0.153 | 1.23 | 0.04 |
| P02768 | ALB | False | True | EV-only | 0.124 | 0.432 | 1.07 | 0.0229 |
| P02787 | TF | False | True | EV-only | 0.216 | 0.236 | 1.29 | 0.0229 |
| P04275 | VWF | True | False | Plasma-only | -1.02 | 0.0117 | -0.243 | 0.641 |
| P05019 | IGF1 | False | True | EV-only |  |  | -2.05 | 0.00388 |
| P08493 | MGP | False | True | EV-only |  |  | -1.7 | 0.00388 |
| P0DJI8 | SAA1 | True | True | Shared | -1.34 | 0.0405 | -1.98 | 0.04 |
| P0DJI9 | SAA2 | True | True | Shared | -1.38 | 0.0347 | -2.22 | 0.0319 |
| P14555 | PLA2G2A | False | True | EV-only |  |  | -3.76 | 0.000244 |
| P15291 | B4GALT1 | False | True | EV-only |  |  | -1.28 | 0.0234 |
| P15502 | ELN | False | True | EV-only |  |  | -1.82 | 0.00484 |
| P18428 | LBP | True | False | Plasma-only | -1.01 | 0.00419 | -1.11 | 0.104 |
| P24592 | IGFBP6 | True | True | Shared | -1.09 | 5.95 x 10^-5^ | -1.6 | 0.0262 |
| P35443 | THBS4 | True | False | Plasma-only | -1.1 | 0.00323 | -0.763 | 0.34 |
| P50281 | MMP14 | False | True | EV-only |  |  | -1.67 | 0.00388 |
| P61769 | B2M | True | False | Plasma-only | -1.07 | 0.00162 | 0.787 | 0.191 |
| P80188 | LCN2 | True | False | Plasma-only | -1.29 | 5.95 x 10^-5^ | -0.885 | 0.207 |
| Q08431 | MFGE8 | False | True | EV-only |  |  | -1.3 | 0.0358 |
| Q12931 | TRAP1 | False | True | EV-only |  |  | -1.2 | 0.0429 |
| Q14213 | EBI3 | False | True | EV-only |  |  | -2.64 | 0.000244 |
| Q14767 | LTBP2 | False | True | EV-only |  |  | -1.53 | 0.00712 |
| Q8N6C8 | LILRA3 | True | False | Plasma-only | -1.11 | 1.62 x 10^-5^ |  |  |
| Q8NBP7 | PCSK9 | False | True | EV-only | -0.292 | 0.244 | -1.39 | 0.0479 |
| Q9BUD6 | SPON2 | False | True | EV-only | -0.441 | 0.126 | -1.46 | 0.0229 |
| Q9H2X0 | CHRD | False | True | EV-only |  |  | -1.28 | 0.0476 |
| Q9UNA0 | ADAMTS5 | False | True | EV-only |  |  | -1.48 | 0.0229 |
